# Accessing abortion in a highly restrictive legal regime: characteristics of women and pregnant people in Malta self-managing their abortion through online telemedicine

**DOI:** 10.1101/2022.10.17.22281157

**Authors:** Andreana Dibben, Isabel Stabile, Rebecca Gomperts, James Kohout

**Author notes:** Correspondence to Andreana Dibben, Department of Social Policy and Social Work, University of Malta, Tal Qroqq, Msida, Malta, MSD2080.

## Abstract

**Objective:** To examine the numbers and characteristics of women and pregnant people in Malta seeking at-home medical abortion using online telemedicine from 2017 to 2021.

**Design:** Population-based study.

**Setting:** Republic of Malta

**Participants:** Between 1 January 2017 and 31 December 2021, 1090 women and pregnant people requested at-home medical abortion through one online telemedicine provider (Women on Web). Mifepristone and misoprostol were shipped to 658 women (60.4% of requests).

**Main Outcome Measures:** The numbers and demographics of persons to which abortion pills were shipped, their reasons for termination of pregnancy, and reasons for requesting medical abortion between January 2017 and December 2021 were analysed. Selected data were compared across different groups.

**Results:** The number of persons in Malta to whom medical abortion pills were shipped increased significantly during the COVID-19 pandemic. Women and pregnant people requesting medical abortion were diverse with respect to age, pregnancy circumstances and reasons for seeking termination. More than half were mothers and over 90% reached out to Women on Web at < 7 weeks. Among those completing a medical abortion, 63% did not use contraception (n=412), and in 30% (n=197) there was contraception failure. The most common reasons for ordering medical abortion pills online were difficulty accessing abortion because of legal restrictions (73%) and abortion pills not being available (45%) in the country.

**Conclusions:** Despite a complete ban on abortion, the number of women and pregnant people residing in Malta completing at-home medical abortions is considerable and has increased since the COVID-19 pandemic.

**Summary Boxes:** *What is already known:* Legal restrictions do not impede women and pregnant people from accessing abortion care but make them liable to criminal prosecution.

*What this study adds:* Since abortion is criminalised in Malta, there are no official statistics related to abortion. This is the first study looking at the number and characteristics of women and pregnant people who access abortion care outside the formal healthcare system in this highly restrictive legal regime. Despite the abortion ban, women and pregnant people in Malta are increasingly accessing telemedicine and self-managing their abortions.

## Introduction

With a population of half a million, Malta is the smallest, southernmost nation state in the EU and its abortion laws are among the most restrictive in the world. Termination of pregnancy (TOP) is illegal under all circumstances.^1^ Medical doctors, fearing prosecution, claim to use the doctrine of double effect to protect the life of at-risk mothers such as in preterm premature rupture of membranes.^2^ Nevertheless, the Criminal Code clearly states that that anyone who intentionally induces a miscarriage is liable to up to three years of imprisonment, while any healthcare professional assisting a woman in procuring an abortion risks up to four years of imprisonment and loss of their license.^3^

The options available for women and pregnant people living in Malta who have a pregnancy they do not want or one that they feel they cannot continue with, have always been limited. Historically, women have either travelled to countries where abortion is legal or remained pregnant, but backstreet abortions were not the norm.^4^ On average, between 2011 and 2019, 56 Maltese residents accessed TOP services in England and Wales.^5^ Travel has never been an easy option; women would need to take time off work, find childcare, fund the abortion, all while keeping the reason for travel a secret from most friends and family. While surmountable for some, these barriers serve as a stark reminder of the disparity in access to essential health care.

Since the COVID-19 pandemic, the number of residents from Malta travelling to the UK to access abortion services declined drastically: 20 in 2020 and only 4 in 2021, even though travelling in 2021 was not as restricted as previously. More women reached out to activist and abortion support groups during the March to May 2020 lockdown period^1^, while there was a significant surge in purchases of abortion pills from Women on Web (WoW) in various European countries including Malta.^6^ This change reflects policy or protocol changes to facilitate access to telemedicine for self-managed abortions implemented in various European countries.^7^ While such changes were obviously not possible in Malta, grassroots organisations stepped in to bridge the gaps. In August 2020, three pro-choice organisations in Malta launched a volunteer-run helpline to facilitate access by providing information on reproductive choices. This Family Planning Advisory Service (FPAS) assisted 479 individuals in its first year alone.^8^

Since 2006, the non-profit organization WoW has provided online telemedicine to women in countries where abortion care is legally restricted.^9^ In 2008, a Maltese newspaper reported that abortion pills were also available to women in Malta^10^ and women have had the option of at-home medical abortion through online telemedicine since then. Between 2013 and 2017, 465 women living in Malta contacted WoW to purchase abortion pills online.^11^ Although this option costs much less than travel abroad, women in Malta having medical abortions at home do so without the reassurance of having doctors they can confide in should any complications arise while exposing themselves to potential criminal liability.

WoW provides an initial online consultation form which is reviewed by a medical doctor. If the gestation is less than 10 weeks and there are no contra-indications, a prescription and package containing mifepristone and misoprostol is dispatched against a donation, which is reduced or waived in cases of financial hardship. Women in Malta usually receive the package within two weeks. A helpdesk team provides real-time instructions and follow up by email. A 10-year study using data from over 26000 women who used the service in countries with highly restrictive legislations worldwide confirmed that self-managed medical abortion is safe and effective.^12^

The aim of this study was to examine the number and characteristics of women in Malta seeking at-home medical abortion through telemedicine from WoW, the main online medical abortion provider. Similar studies have been conducted in Ireland, Northern Ireland, Hungary, Germany, and Italy in recent years.^13–18^ Smaller alternative services e.g., Women Help Women, are also available, such that this cohort does not represent the entire population obtaining medical abortion pills online.

## Methods

We retrieved all requests to WoW for medical abortion from women in Malta who completed an online consultation between January 2017 and December 2021. This date range was chosen because the consultation forms that had been developed over time had become constant such that almost complete data were available. The consultation form includes information about demographics, medical and pregnancy history, and health status. Women also select the circumstances of their pregnancy, the reasons for needing an abortion and their reasons to order medical abortion pills online. All the questions have predefined answers and women can choose as many responses as they wish. As nationality is not included, we cannot distinguish between citizens and non-citizens, although it is estimated that 20% of the population are non-Maltese citizens.^19^

De-identified data were provided by WoW and analysed using Microsoft SPSS. Women had consented to anonymised use of their data for research purposes. The Faculty for Social Wellbeing Research Ethics Committee, University of Malta reviewed this study.

We analysed the number of women to whom medication was shipped between 2017 and 2021. For all five years combined, we examined the age distribution, number of children, weeks of gestation, circumstances of pregnancy, reasons for TOP and reasons for accessing telemedicine. We compared data before and during COVID for women greater or less than 20 years and those with or without children. The Chi square test was used to investigate the association between categorical variables. Findings were considered statistically significant at a *p* value of < 0.05. Most women did not complete the follow-up evaluation, which was therefore not included in the analysis.

### Patient and public involvement statement

Although this analysis did not involve patients in its design, management or reporting, the research questions were informed by the experiences of women and pregnant people in Malta who rely on WoW to access abortion. Some preliminary results were published in a Maltese newspaper^20^ and presented in an academic conference.^21^ Feedback from public and academic peers was considered.

## Results

The number of women living in Malta seeking medical abortion through WoW increased steadily from 78 in 2017 to 509 in 2021, a more than sixfold increase. Between January 2017 and December 2021, 1090 women contacted WoW to request medical abortion and medication was shipped to 658 women (60.4% of requests). The remainder were cancelled either because they chose to continue the pregnancy, experienced a miscarriage, or decided on travel to terminate their pregnancy abroad.

Figure 1 shows the number of abortion packages shipped per year. There was a 33% decrease in the number of shipments between 2017 and 2018, but a 45% increase between 2018 and 2019, a 108% increase between 2019 and 2020, and a 65% increase between 2020 and 2021. The sharpest increase was observed in 2020, corresponding to the onset of the COVID-19 pandemic. Shipments increased by 47% between March and June 2019 and the same months of 2020 (27 and 51, respectively).

**Figure 1.**
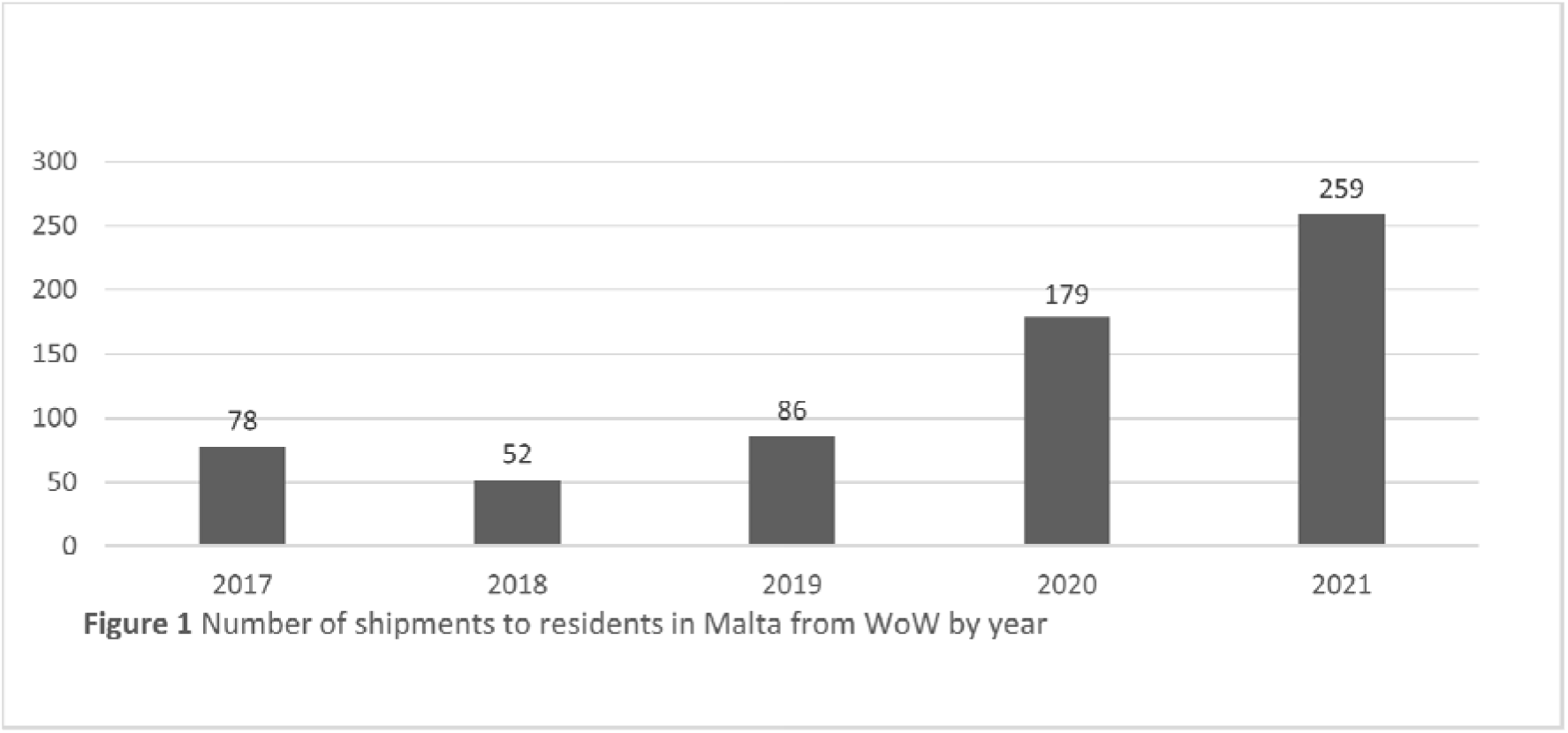
Number of shipments to residents in Malta from WoW by year

Table 1 shows the characteristics of women and pregnant people to whom abortion medication was shipped between 2017 and 2021. It further differentiates between before and during the COVID-19 pandemic. The mean age was 29.3 years, but the cohort included women from all reproductive age groups. Just over half (53.7%) were between 25 and 34 years old, nearly a quarter (22%) were women 35 years and over, and another quarter (24.3%) were under 25. The majority (52%) were mothers with a mean number of 1.72 children while almost a quarter (24%) had two or more children. 92% made the request at a gestational age of less than seven weeks.

**Table 1.**
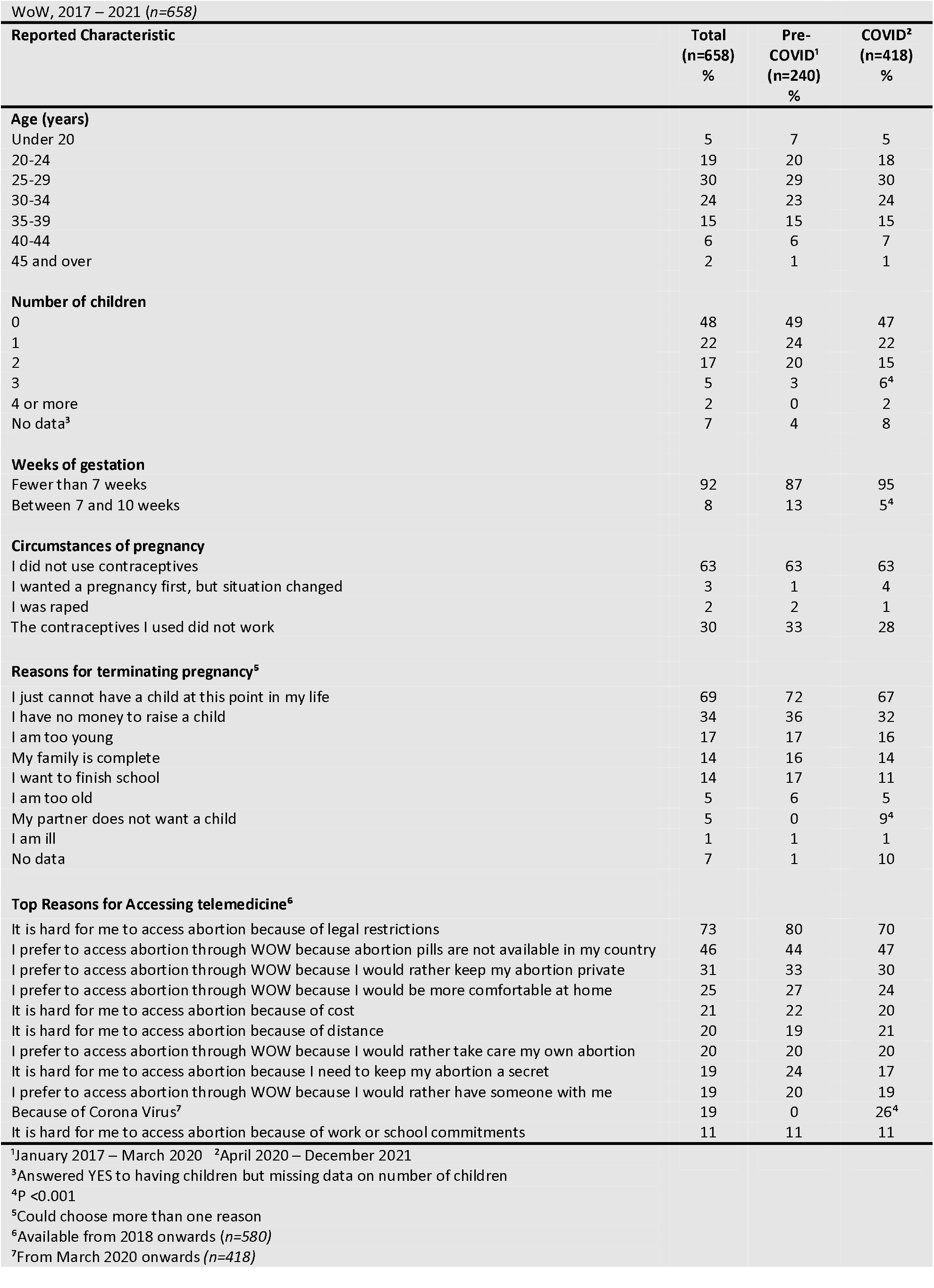
Characteristics of women and pregnant people living in Malta to whom abortion packages were shipped through

| WoW, 2017 – 2021 (*n*=658) |  |  |  |
| --- | --- | --- | --- |
| Reported Characteristic | Total<br>(*n*=658)<br>% | Pre-COVID<sup>1</sup><br>(*n*=240)<br>% | COVID<sup>2</sup><br>(*n*=418)<br>% |
| **Age (years)** |  |  |  |
| Under 20 | 5 | 7 | 5 |
| 20-24 | 19 | 20 | 18 |
| 25-29 | 30 | 29 | 30 |
| 30-34 | 24 | 23 | 24 |
| 35-39 | 15 | 15 | 15 |
| 40-44 | 6 | 6 | 7 |
| 45 and over | 2 | 1 | 1 |
| **Number of children** |  |  |  |
| 0 | 48 | 49 | 47 |
| 1 | 22 | 24 | 22 |
| 2 | 17 | 20 | 15 |
| 3 | 5 | 3 | 6<sup>4</sup> |
| 4 or more | 2 | 0 | 2 |
| No data<sup>3</sup> | 7 | 4 | 8 |
| **Weeks of gestation** |  |  |  |
| Fewer than 7 weeks | 92 | 87 | 95 |
| Between 7 and 10 weeks | 8 | 13 | 5<sup>4</sup> |
| **Circumstances of pregnancy** |  |  |  |
| I did not use contraceptives | 63 | 63 | 63 |
| I wanted a pregnancy first, but situation changed | 3 | 1 | 4 |
| I was raped | 2 | 2 | 1 |
| The contraceptives I used did not work | 30 | 33 | 28 |
| **Reasons for terminating pregnancy<sup>5</sup>** |  |  |  |
| I just cannot have a child at this point in my life | 69 | 72 | 67 |
| I have no money to raise a child | 34 | 36 | 32 |
| I am too young | 17 | 17 | 16 |
| My family is complete | 14 | 16 | 14 |
| I want to finish school | 14 | 17 | 11 |
| I am too old | 5 | 6 | 5 |
| My partner does not want a child | 5 | 0 | 9<sup>4</sup> |
| I am ill | 1 | 1 | 1 |
| No data | 7 | 1 | 10 |
| **Top Reasons for Accessing telemedicine<sup>6</sup>** |  |  |  |
| It is hard for me to access abortion because of legal restrictions | 73 | 80 | 70 |
| I prefer to access abortion through WOW because abortion pills are not available in my country | 46 | 44 | 47 |
| I prefer to access abortion through WOW because I would rather keep my abortion private | 31 | 33 | 30 |
| I prefer to access abortion through WOW because I would be more comfortable at home | 25 | 27 | 24 |
| It is hard for me to access abortion because of cost | 21 | 22 | 20 |
| It is hard for me to access abortion because of distance | 20 | 19 | 21 |
| I prefer to access abortion through WOW because I would rather take care my own abortion | 20 | 20 | 20 |
| It is hard for me to access abortion because I need to keep my abortion a secret | 19 | 24 | 17 |
| I prefer to access abortion through WOW because I would rather have someone with me | 19 | 20 | 19 |
| Because of Corona Virus<sup>7</sup> | 19 | 0 | 26<sup>4</sup> |
| It is hard for me to access abortion because of work or school commitments | 11 | 11 | 11 |
| <sup>1</sup>January 2017 – March 2020 <sup>2</sup>April 2020 – December 2021 |  |  |  |
| <sup>3</sup>Answered YES to having children but missing data on number of children |  |  |  |
| <sup>4</sup>P <0.001 |  |  |  |
| <sup>5</sup>Could choose more than one reason |  |  |  |
| <sup>6</sup>Available from 2018 onwards (*n*=580) |  |  |  |
| <sup>7</sup>From March 2020 onwards (*n*=418) |  |  |  |

When describing the circumstances of their pregnancy, most women were either not using contraception (63%) or had experienced failure of contraception (30%). The most common reasons cited for the termination of pregnancy were not being able to have a child at this point in their life (69%) and having no money to raise a child (34%). The most common reasons for accessing telemedicine were legal restrictions (73%) and unavailability of medical abortion pills (46%).

There were some significant differences in the characteristics of women accessing abortion through telemedicine in the period before compared with during COVID-19. Significantly more women with three or more children accessed abortion during the COVID-19 period (n=33) compared to before the pandemic (n=7) (*p* < 0.001). Women were also more likely to purchase abortion pills earlier in the pregnancy during the pandemic (*p* < 0.001). During COVID-19 more women responded ‘My partner does not want a child’ as a reason for terminating the pregnancy (n=38 compared to n=0). (*p* < 0.001)

Table 2 compares the circumstances of pregnancy and reasons for termination selected by women with and without children. Having children seems to influence the reasons for seeking an abortion since mothers were significantly more likely to select ‘I am too old’ and ‘My family is complete’ than women without children. Those without children were more likely to want to finish school or select ‘I have no money to raise a child’ as a reason. Women who were mothers were also less likely to be using contraception than those without children (70% compared to 60%) and women without children were more likely to experience contraception failure than those who were mothers (36% compared to 25%).

**Table 2.** Comparison of selected characteristics of persons with and without children to whom abortion packages were shipped through WoW, 2017 – 2021

| Selected Characteristic | Children |  |
| --- | --- | --- |
|  | Yes<br>(n=344) | No<br>(n=314) |
| **Circumstances of pregnancy (p = 0.036)** |  |  |
| I did not use contraceptives | 70 (241) | 60 (188) |
| I wanted a pregnancy first, but situation changed | 3 (11) | 2 (6) |
| I was raped | 2 (6) | 2 (6) |
| The contraceptives I used did not work | 25 (86) | 36 (113) |
| **Reasons for terminating pregnancy (p < 0.001)** |  |  |
| I just cannot have a child at this point in my life | 60 (206) | 72 (226) |
| I have no money to raise a child | 29 (98) | 36 (112) |
| My family is complete | 27 (93) | 3 (8) |
| I want to finish school | 5 (16) | 20 (62) |
| I am too old | 9 (32) | 2 (5) |
| My partner does not want a child | 5 (17) | 5 (16) |
| All data are given as a percentage (n) |  |  |

The reasons that younger persons gave for accessing abortion also differed from the older cohort (Table 3). Women younger than 20 were significantly more likely to state that they wanted to finish school (21% versus 11%), that they have no money to raise a child (59% versus 32%) and were less likely to state that their family was complete (3% versus 15%) (*p* < 0.001). Teenagers more frequently selected the need to keep abortion a secret, the cost, and school and work commitments and fewer reported having a partner or friend with them during the process (*p* = 0.041). Teenagers and adults did not differ significantly in terms of gestation when requesting abortion pills.

**Table 3.**
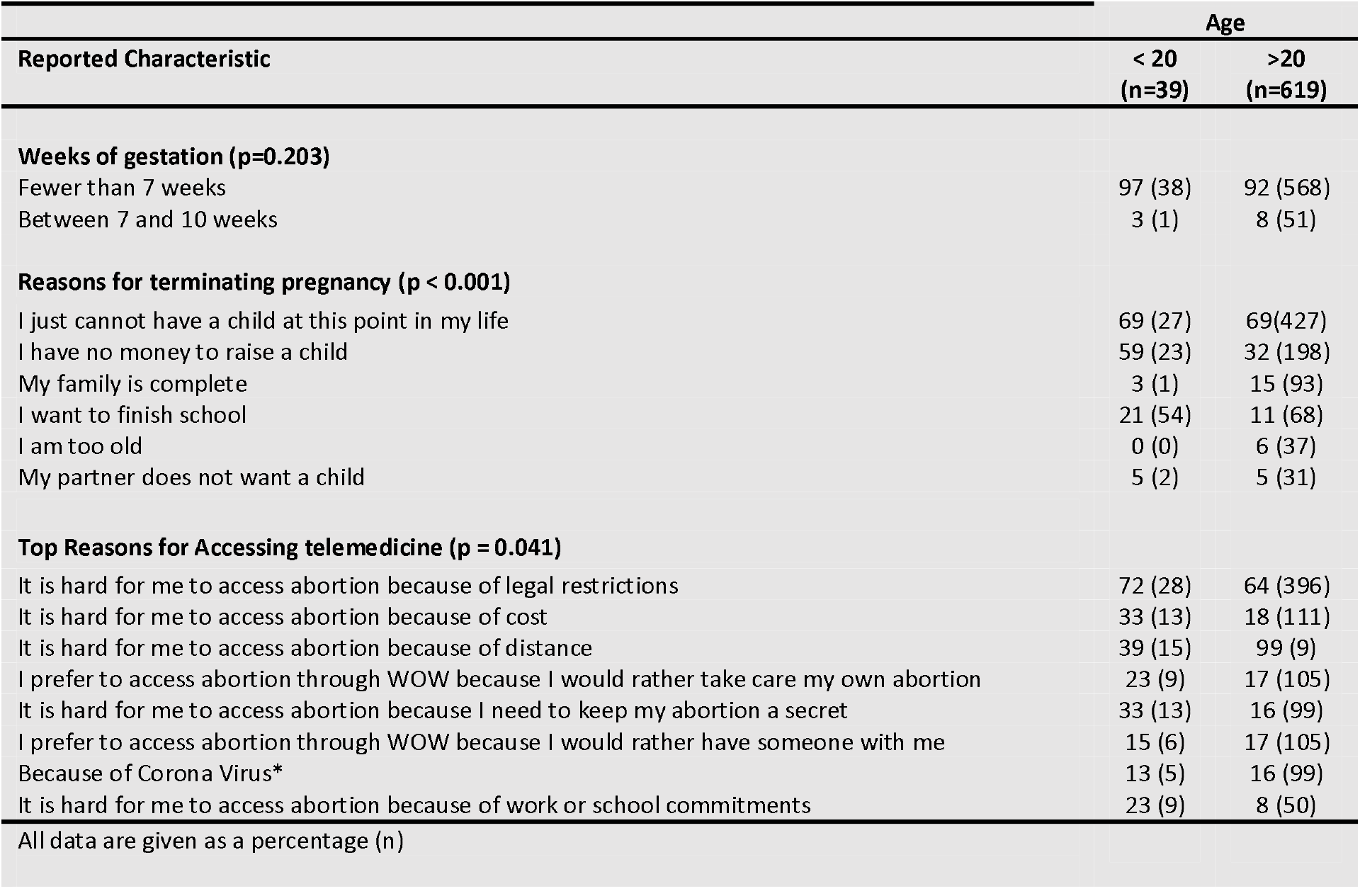
Comparison of selected characteristics of persons over and under 20 to whom abortion packages were shipped through WoW, 2017 – 2021

## Discussion

The most common reasons cited by women and pregnant people in Malta for accessing abortion through telemedicine were legal restrictions and the lack of availability of abortion pills, further evidence that legal restrictions do not stop abortions taking place. Especially since the COVID-19 pandemic, abortion care by women in Malta has shifted from travelling abroad to countries where abortion is legal to self-managing medical abortion after purchasing abortion pills online. The significant and increasing demand for self-managed abortions is congruent with developments in other regions where abortion care is restricted. A literature analysis of studies in Latin American countries found that women value the privacy that medical abortion allows as well as the possibility of having someone with them during the process. They perceive it as less painful, safer, more practical, less expensive, and more natural than other abortion methods.^22^

The implication of the increasing demand for at-home medical abortion is that more women in Malta are now breaking the law to access abortion, increasing their need for secrecy due to potential criminal prosecution. In a Guardian interview^23^, one woman described her fear when a local politician threatened to prosecute women who had sought abortion care in other countries. While this fear was unfounded, since women who seek abortion care outside Maltese jurisdiction cannot be prosecuted on their return, this is not the case for women who manage their own abortions within the country. As indicated in the Latin American study, the disadvantage of medical abortion is the possible need to seek medical care where they may be reported to the police for having an abortion. In another abortion story^24^, the woman recounted experiencing prolonged bleeding, pain, and disorientation and her reluctance to seek medical care due to the risk of prosecution.

In a context where abortion is not legally available through formal healthcare services, women who seek abortion care through telemedicine are diverse in terms of age and circumstances. At 29.3, the median age is very similar to that found in studies in countries such as Italy, Hungary, and Ireland where the mean age was 30. Teenagers constitute 5% of women receiving abortion pills, which is similar to Ireland (4.6%), higher than Hungary (2.9%) but less than Italy (7.8% calculated at 18 or younger). Although Malta has the lowest fertility rate in the EU, it has a higher-than-average teenage birth rate (11.9 births per 1000 girls aged 15 to 19 in 2019, compared to the EU average of 8.9). This high rate is partly due to the total ban on abortion, considering that teenagers face more barriers to access abortion services^25^. Indeed, the findings from this study highlight how teenagers are more vulnerable since they often face economic and social difficulties and lack the support of a family member or partner. Similar findings were reported in studies of Italian and German women seeking abortion through WoW^16 17^.

Contrary to the prevailing stereotype that most women who terminate their pregnancies are young and promiscuous, most women seeking abortion through telemedicine in Malta were in their late twenties or early thirties and already mothers, and their main reason for termination is that they feel their family is complete or they cannot cope with another child. Notably, among this group, the lack of use of contraception rather than contraception failure was the main reason why women became pregnant. In a recent study, the rate of unintended pregnancies was reported at 23%, mainly due to not using contraception at all (20%) or due to relying on less reliable methods such as withdrawal, natural family planning, or condoms.^26^ Malta lacks a family planning policy or public family planning clinics and most contraception is available upon payment from private health services.^25^ The increase in women with more children accessing abortion during COVID-19 might be indicative of the financial and care-giving hardships that women with larger families faced during the pandemic. The fear and uncertainty that the pandemic brought with it might have led to more partners pressuring women to terminate their pregnancy.

In conclusion, since 2018, the practice of abortion care among women and pregnant people in Malta has steadily shifted to accessing at-home medical abortion through telemedicine, exacerbated by travel restrictions during the COVID-19 pandemic and by the local provision of voluntary services such as FPAS providing information and support.

Although at-home medical TOP is medically safe, women and pregnant people in Malta who access WoW services risk criminal prosecution, drawing attention to the urgent need to decriminalise abortion to make it safer and to address the inequities in accessing reproductive health services, as recommended by the WHO.^27^

This study further highlights the need for better family planning services to reduce barriers to access especially among women with children, and the promotion of more reliable methods of contraception. It further indicates that teenagers seeking self-managed abortion might be in a more vulnerable situation since they are more likely to lack support.

## Data Availability

All data produced in the present study are available upon reasonable request to the authors

## Acknowledgements

The authors thank Prof Liberato Camilleri, University of Malta for assistance with statistical data.

## Contributors

AD and IS designed the study. RG supplied the data. JK performed the statistical analysis. AD and IS wrote the initial manuscript, interpreted the data, and revised subsequent drafts. All read and approved the final draft of the manuscript for submission and accept responsibility for the paper as published. IS is the guarantor.

## Copyright/License for Publication

The Corresponding Author has the right to grant on behalf of all authors and does grant on behalf of all authors, a worldwide licence to the Publishers and its licensees in perpetuity, in all forms, formats and media (whether known now or created in the future), to i) publish, reproduce, distribute, display and store the Contribution, ii) translate the Contribution into other languages, create adaptations, reprints, include within collections and create summaries, extracts and/or, abstracts of the Contribution, iii) create any other derivative work(s) based on the Contribution, iv) to exploit all subsidiary rights in the Contribution, v) the inclusion of electronic links from the Contribution to third party material where-ever it may be located; and, vi) licence any third party to do any or all of the above.

## Ethics Approval

The Faculty for Social Wellbeing Research Ethics Committee, University of Malta reviewed this study..

## Transparency Statement

Isabel Stabile, the manuscript’s guarantor affirms that the manuscript is an honest, accurate, and transparent account of the study being reported; that no important aspects of the study have been omitted; and that any discrepancies from the study as originally planned (and, if relevant, registered) have been explained.

## Data Sharing

The authors confirm that they are willing to share all anonymised data on which the analysis, results, and conclusions reported in the paper are based.

